# Maximum entropy method for estimating the reproduction number: An investigation for COVID-19 in China

**DOI:** 10.1101/2020.03.14.20035659

**Authors:** Yong Tao

## Abstract

The key parameter that characterizes the transmissibility of a disease is the reproduction number *R*. If it exceeds 1, the number of incident cases will inevitably grow over time, and a large epidemic is possible. To prevent the expansion of an epidemic, *R* must be reduced to a level below 1. To estimate the reproduction number, the probability distribution function of the generation interval of an infectious disease is required to be available; however, this distribution is often unknown. In this letter, given the incomplete information for the generation interval, we propose a maximum entropy method to estimate the reproduction number. Based on this method, given the mean value and variance of the generation interval, we first determine its probability distribution function and in turn estimate the real-time values of reproduction number of COVID-19 in China. By applying these estimated reproduction numbers into the susceptible-infectious-removed epidemic model, we simulate the evolutionary track of the epidemic in China, which is well in accordance with that of the real incident cases. The simulation results predict that China’s epidemic will gradually tend to disappear by May 2020 if the quarantine measures can continue to be executed.

---

In December, 2019, a cluster of pneumonia cases in Wuhan, China was caused by a novel coronavirus, the COVID-19 [1-4]. At first the local governments did not take effective measures which leaded to local people not paying enough attention to the risk. However, with the epidemic in Wuhan further expanding, the Chinese Government started to take emergency actions to lock down the Wuhan city on January 23, 2020. Despite this, the epidemic still spread throughout the entire country. By March 12, 2020, controlling the spread of the epidemic has become a global challenge. One of the key parameters in epidemic models is the basic reproduction number *R*_0_, defined as the number of secondary infections that arise from a typical primary case in a completely susceptible population [5]. As an infection is spreading through a population, it is more convenient to work with an effective reproduction number *R*_*t*_, which is defined as the number of secondary infections that arise from a typical primary case [5]. The magnitude of *R*_*t*_ is a useful indicator for evaluating the risk of an infectious disease and the validity of controlling the epidemic. If *R*_*t*_ exceeds 1, the number of incident cases will inevitably grow over time, and a large epidemic is possible. To prevent the expansion of an epidemic, *R*_*t*_ must be reduced to a level below 1. Using the parameter *R*_*t*_, one can establish the susceptible-infectious-removed (SIR) epidemic model as below [6-8]:

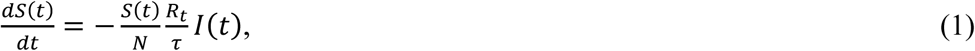

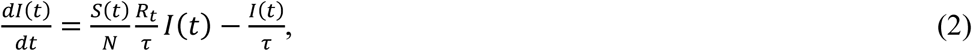

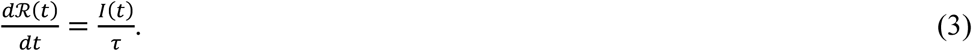

where *S*(*t*), *I*(*t*) and *R*(*t*) are the number of susceptible, infectious, and removed (including recovered and death) individuals at time *t*; *τ* denotes the generation interval that is the time from infection of an individual to the infection of a secondary case by that individual, namely, the “contagion period” of an infection [5]. The generation interval *τ* should be a random variable. If we denote the number of populations by *N*, we have:

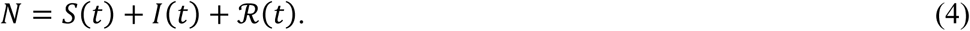

If *R*_*t*_ and *τ* of an epidemic are known, one can employ the SIR model (1)-(4) to simulate the evolutionary track of this epidemic. To estimate the reproduction number *R*_*t*_, the probability distribution function of the generation interval of an infectious disease, *p*(*τ*), is required to be available [5, 9-12]; however, this distribution is often unknown. In the existing literature, many scholars used exponential distribution [5], normal distribution [5], Weibull distribution [9, 10], and Gamma distribution [3, 12] to approximate *p*(*τ*). Theoretically, to use these distributions to approximate *p*(*τ*), one needs to know enough information about symptom onsets of all cases, namely, large sample cases for *τ*. Regarding the incomplete information, one also applied the Monte-Carlo method [4] and Bayesian statistical inference [11] to estimating *p*(*τ*). However, thus far, there is scant literature to discuss the potential application of the maximum entropy method (MaxEnt) [13-15] in estimating the reproduction number. Our letter fills this gap. In the statistical inference, MaxEnt is a powerful tool of predicting probability distributions. The main idea of MaxEnt is to estimate a target probability distribution by finding the probability distribution of maximum entropy, subject to a set of constraints that represent our incomplete information for the target distribution [13]. Due to the advanced predictive capacity, MaxEnt has been widely applied in thermodynamics [13], economics [16-19], artificial intelligence [20-21], and ecology [22-26].

In this letter, we apply the MaxEnt to determining the function shape of *p*(*τ*). Before doing so, we first introduce the relationship between *R*_*t*_ and *p*(*τ*). Here, we adopt Wallinga and Lipsitch’s method [5] for deriving the reproduction number. By both authors’ method, the number of infectious individuals at time *t* can be written as [5]:

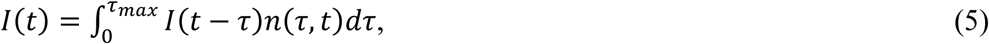

where *n*(*τ, t*) denotes the number of cases infected by a *τ*-day infectious individual at time t. Here *τ*_*max*_ denotes the maximum symptom duration. Wallinga and Lipsitch assumed [5] *τ*_*max*_ = +∞. To make the model more realistic, we assume that *τ*_*max*_ is a finite number. Thus, the reproduction number *R*_*t*_ can be defined as [5]:

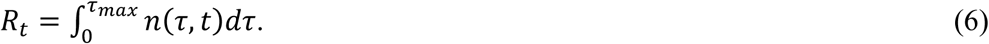

Let us order

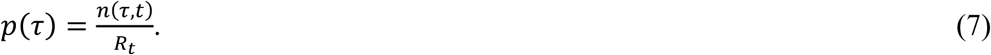

Substituting equation (7) into equation (6) yields:

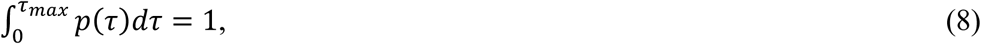

where *p*(*τ*) is the probability distribution function of the generation interval *τ* [5].

Using equation (8), the mean value of the generation interval can be written as:

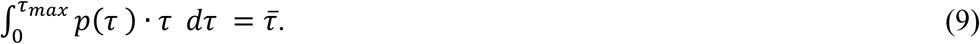

If the mean value 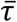 is known, then by using equation (8) one can obtain the variance of the generation interval:

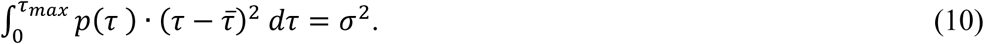

Substituting equation (7) into equation (5) we finally obtain:

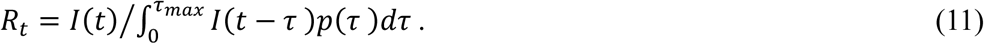

Equation (11) is the basic formula for calculating the reproduction number. If *I*(*t*) and *p*(*τ*) are known, one can calculate the reproduction number by using equation (11). Generally speaking, the number of infectious individuals *I*(*t*) is reported for each day, while the function shape of *p*(*τ*) is unknown. Therefore, many scholars used exponential distribution [5], normal distribution [5], Weibull distribution [9, 10] and Gamma distribution [3, 12] to approximate *p*(*τ*). To do so, one needs to collect enough information of *τ*, which requires examining a large number of cases. From a practical point of view, it is easier to collect a sample set of cases (at least 30 samples) to calculate the approximate estimates of the mean value 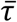 and the variance *σ*^2^. Given the approximate estimates of 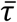 and *σ*^2^ as the prior information, we maximize the information entropy of the generation interval *τ* to infer the function shape of the probability distribution *p*(*τ*). This is the basic idea of the MaxEnt, which agrees with everything that is known, but avoids assuming anything that is not known [15]. The resulting statistical inference gives the least biased predictions of the shapes of probability distributions consistent with prior knowledge [23].

Now we apply the MaxEnt to determining the probability distribution function *p*(*τ*). To this end, we assume that the mean value 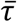 and the variance *σ*^2^ of the generation interval *τ* are known. By equation (8), we define the information entropy of the generation interval *τ* as:

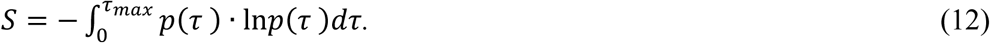

Because we only know the mean value 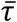 and the variance *σ*^2^, maximizing the information entropy (12) should yield:

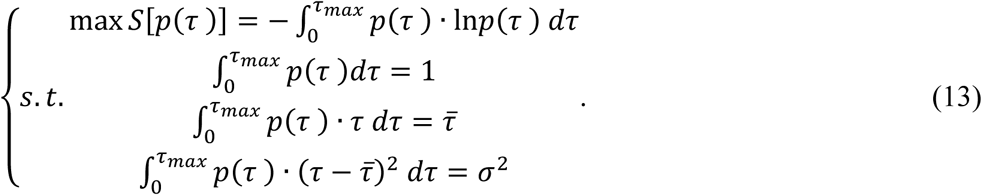

To solve the optimal problem (13), we construct the Lagrange function:

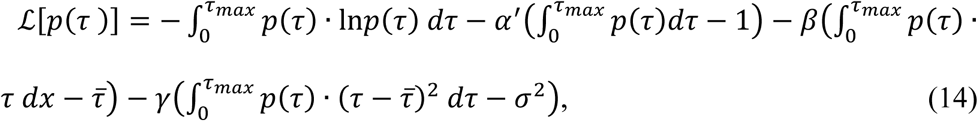

where *α*′, *β*, and *γ* are Lagrange multipliers.

Plugging equation (14) into the functional derivative δℒ[*p*(*τ*)]/δ*p*(*τ*) = 0 we get the optimal solution:

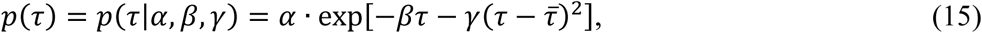

where *α* = exp(−*α*′ − 1).

Theoretically, substituting equation (15) into equations (8), (9), and (10) one can calculate the values of *α, β*, and *γ*. However, it is difficult to obtain the analytic results of integrals (8), (9) and (10). To do numerical calculation for equations (8), (9), and (10), we assume that *p*(*τ*) quickly tends to 0 as *τ ≫* 1. The validity of this assumption can be justified by checking equation (15); therefore, equations (8), (9) and (10) can be written as:

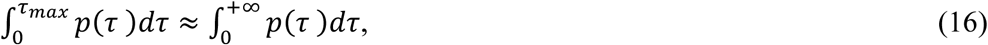

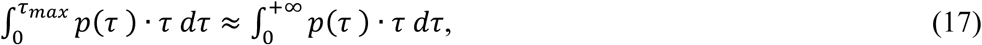

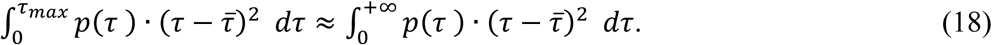

Based on equations (16), (17) and (18), we propose a numerical method to calculate the approximate values of *α, β*, and *γ*. To this end, let us order:

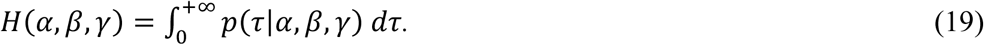

The partial derivatives of equation(19) with respect to *β* and *γ* yield:

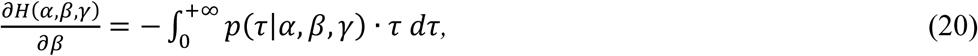

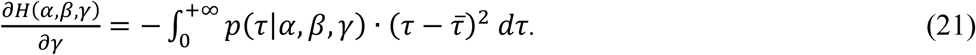

Using equations (19), (20) and (21), equations (8), (9) and (10) can be rewritten in the form:

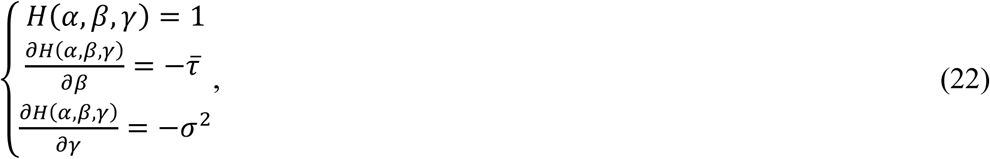

where we have used the approximations (16), (17) and (18).

By solving equation (22), one can obtain the approximate values of *α, β*, and *γ*. Substituting equation (15) into equation (19) we have

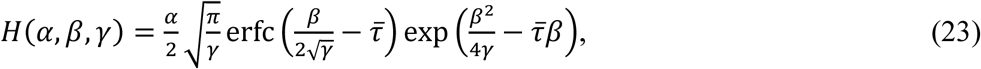

where 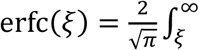 exp(−*x*^2^) *dx* denotes the error function.

By equation (23) it is easy to get:

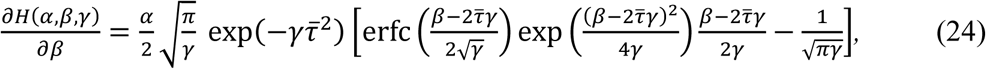

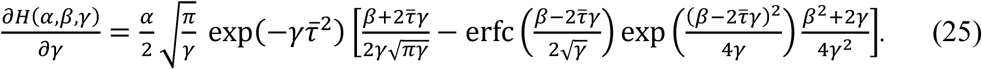

Solving equation (22) is equivalent to minimizing the following function:

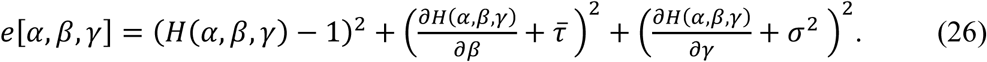

Here we employ the Matlab software to depict *e*[*α, β, γ*] as a 100 × 100 × 100 lattice-point matrix, where the lattice spacing is 0.01. Given the accuracy of 0.01, by inputting the observed values of 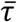 and *σ*^2^, we calculate *α, β*, and *γ*. We first apply equation (26) to the SARS epidemic in Singapore in 2003, where 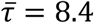 (days) and *σ* = 3.8 (days) [9]. Substituting both observed values into equation (26), we seek the lattice-point minimizing equation (26) as below:

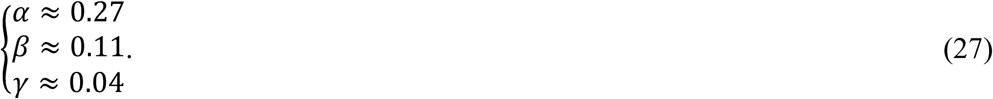

Substituting equation (27) into equation (15) we have:

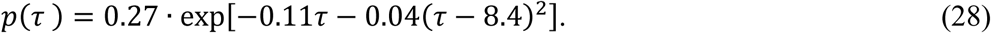

The shape of equation (28) is showed in the Figure 1, see the blue curve. It is well in accordance with the sample data of generation interval of the SARS in Singapore in 2003, see the red histogram in Figure 1. This result supports the validity of the MaxEnt. The latest clinical research [1] showed that the generation interval of COVID-19 is similar to that of SARS. Therefore, we assume that the generation interval of COVID-19 shares the same probability function shape as that of SARS. This assumption was also adopted by Wu et al [4]. Based on this assumption, we apply equation (28) to estimating the reproduction number of COVID-19 in China. To this end, substituting equation (28) into equation (11) we have:

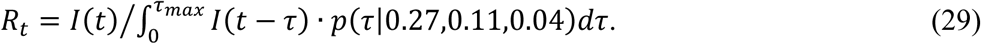

**Figure 1.**
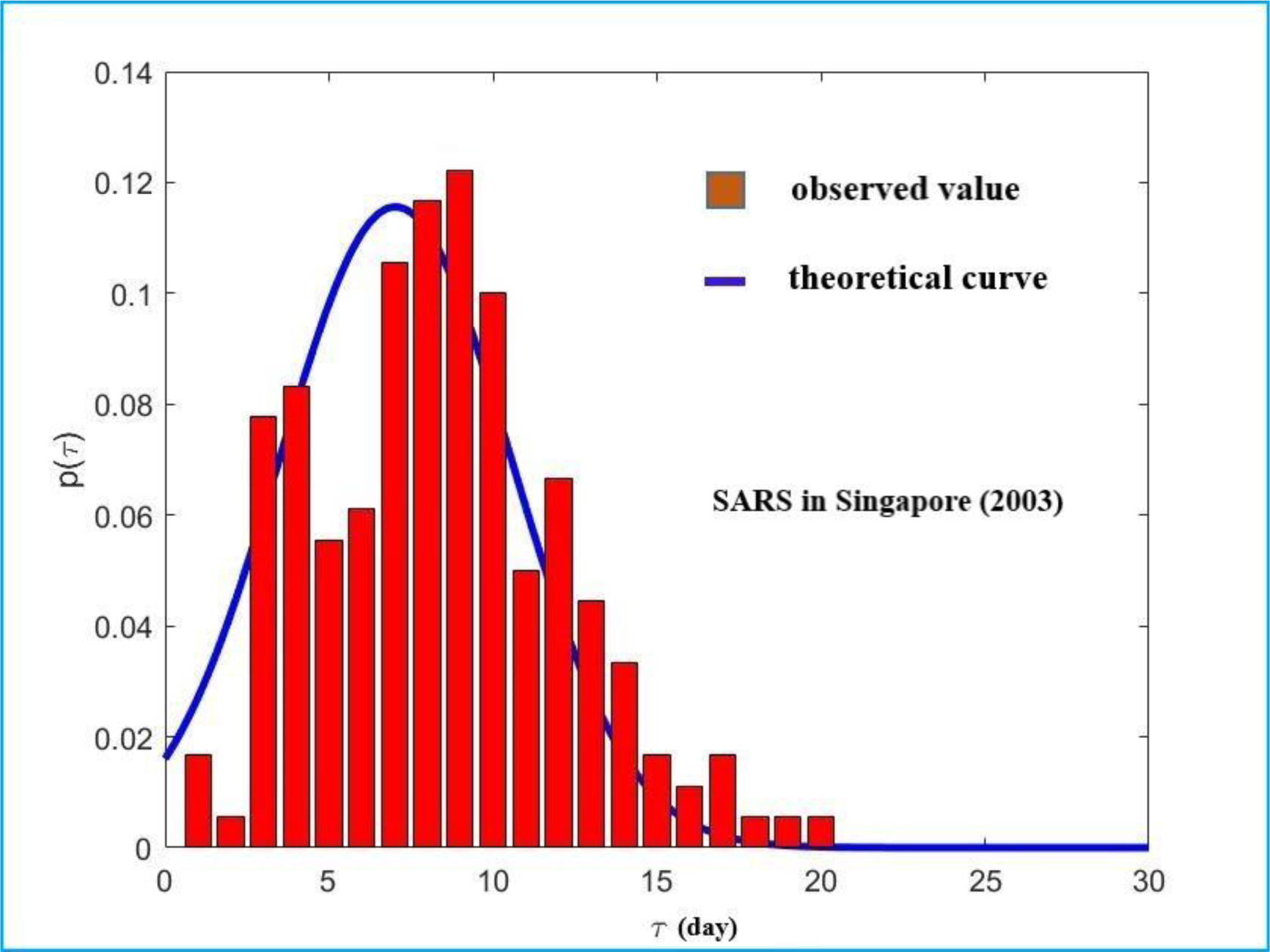
The function shape of equation (28) is showed by a blue curve. The sample data of generation interval of the SARS in Singapore (2003) is showed by a red histogram, and the data resource refers to [9].

Because the observed data of *I*(*t*) was reported for each day, we denote the unit of time t by “day”. To calculate *R*_*t*_ by using equation (29), we need to rewrite the integral (29) as a summation formula. To do so, by Figure 1 we observe *p*(*τ* = 14) *≈* 0. By equations (16)-(18), this means *τ*_*max*_ *≈* 14. Therefore, the maximum generation interval of COVID-19 can be approximately denoted by 14. Based on this setting, equation (29) can be rewritten in the form:

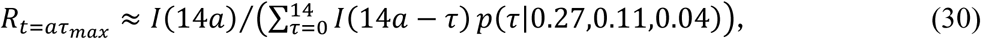

where *a* = 1,2, … denote the ordinal number of the period and *τ*_*max*_ = 14. Without loss of generality, in equation (30) we have approximately identified the reproduction number of the last day of a period as the reproduction number of this period. From the perspective of entire time span of an epidemic, this approximation satisfies the spirit of statistical mean-field method.

By using equation (30) we can report the reproduction number every 14 days. Before doing so, we first determine the starting point of each contagion period for COVID-19 in China. According to the report of China CDC [2], January 8, 2020 was considered as the last day of a contagion period in Wuhan, China; therefore, we mark January 9, 2020 as the first day of the next contagion period. Based on this setting, the first period we report is from January 9, 2020 to January 22, 2020. In fact, this setting agrees with the date of locking down the Wuhan city, January 23, 2020. Here, we have collected the national-level data of the accumulative infected, recovered, and death cases of China’s epidemic from January 10, 2020 to March 4, 2020, see Figure 2. By using the data in Figure 2, it is easy to calculate the number of real-time infected cases, *I*(*t*), in China for each day. This result has been shown in Figure 3. Using the data in Figure 3, we can report the reproduction number every 14 days by using equation (30). The results have been listed in Table 1. The first period (from January 9, 2020 to January 22, 2020) can be regarded as a free propagation stage of COVID-19 because most Chinese people were aware of the outbreak of COVID-19 after January 21, 2020 and local governments did not take effective measures to control the epidemic during this period. Unfortunately, the data in the first period is very incomplete. By contrast, the reported infected cases on January 10 (41 cases) and January 22 (571 cases) can be roughly used. Consider that the first period is a free propagation stage, we use both data to approximately restore real-time data of this period by the exponential growth formula *57*1 = 41. exp(12. *r*), where *r* denotes the growth rate. Using the restored data, the estimated value of the reproduction number for the first period is calculated to be 3.7069, see Table 1, which implies that the intensity of free transmission of COVID-19 is quite high. The World Health Organization announced [27], up to March 12, 2020, the COVID-19 had spread to 118 countries. The rapid worldwide spread of COVID-19 is an evidence for supporting our calculation. After January 22, 2020, Chinese Government started to take emergency actions to lock down the Wuhan city, and quickly performed different quarantine measures in every provinces. The powerful quarantine measures substantially reduce the contagion probability among individuals. Therefore, the subsequent periods no longer belong to free transmission. For these periods, the results of the reproduction number have been listed in Table 1. Due to the quarantine measures, the reproduction number for the second period (from January 23, 2020 to February 5, 2020) has been reduced to 3.122 with the reduction amplitude being 15.78%. For the third period (from February 6, 2020 to February 19, 2020), the reproduction number has remarkably been reduced to 1.2114, which is close to 1. This implies that the epidemic has been effectively controlled. It should be pointed out that the last day of the third period (February 19) is just the turning point of the epidemic, refer to Figure 3; therefore, the real data supports our calculations for the reproduction number. As the epidemic come to the fourth period (from February 20, 2020 to March 4, 2020), the reproduction number is eventually reduced to 0.6028, a level below 1. In this sense, China’s quarantine measures have obtained a preliminary success.

**Table 1:** The reproduction number for each period (2020)

| Period number $a$ | Period interval | $R_t$ | Amplitude reduction |
| --- | --- | --- | --- |
| 1 | 9-Jan to 22-Jan | 3.7069 | — |
| 2 | 23-Jan to 5-Feb | 3.1220 | 15.78% |
| 3 | 6-Feb to 19-Feb | 1.2114 | 61.20% |
| 4 | 20-Feb to 4-Mar | 0.6028 | 50.24% |

**Figure 2.**
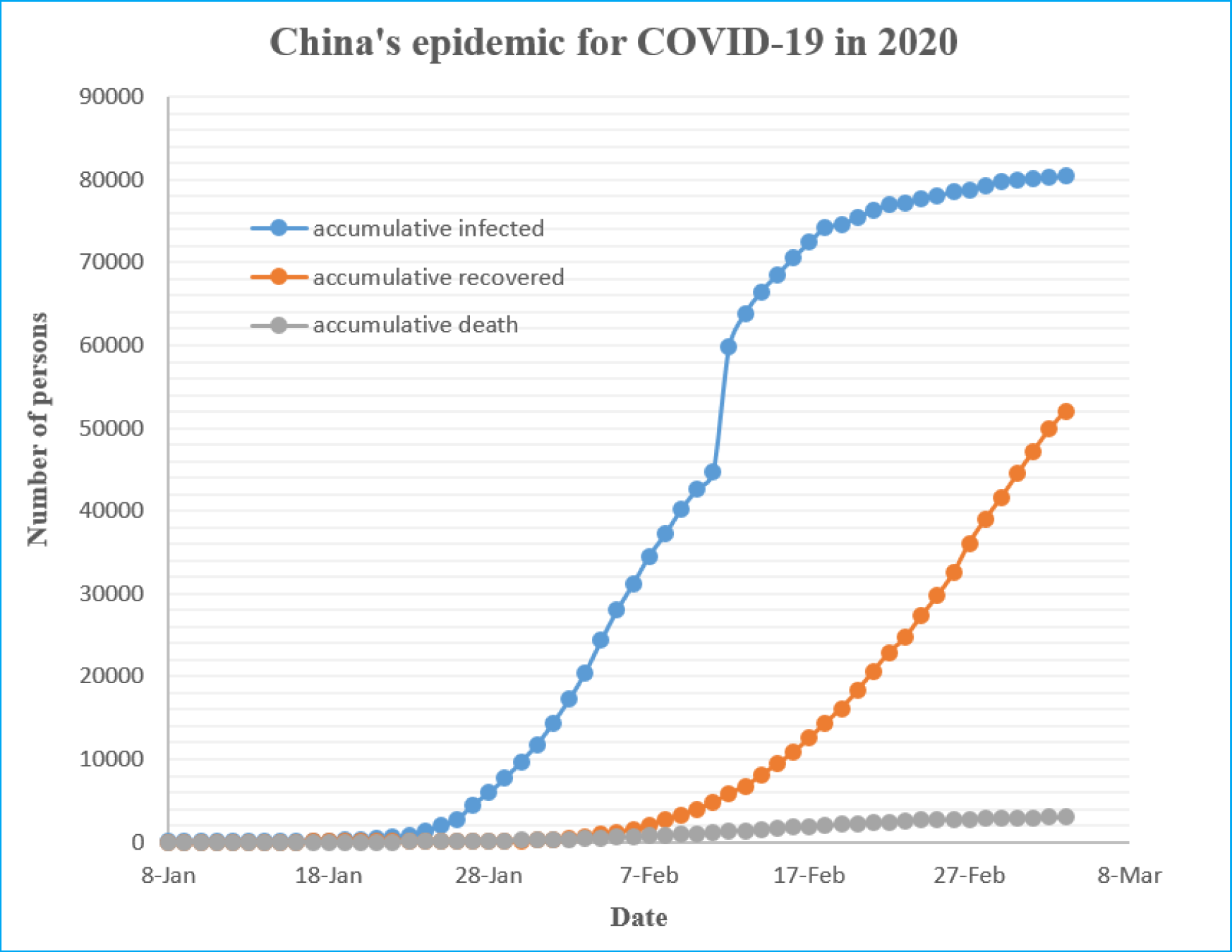
National-level data of the accumulative infected, recovered, and death cases of China’s epidemic from January 10, 2020 to March 4, 2020. The data of January 8 and 9 is simply assumed to be same as that of January 10. **Data resource:** https://voice.baidu.com/act/newpneumonia/newpneumonia/?from=osari_pc_3

**Figure 3.**
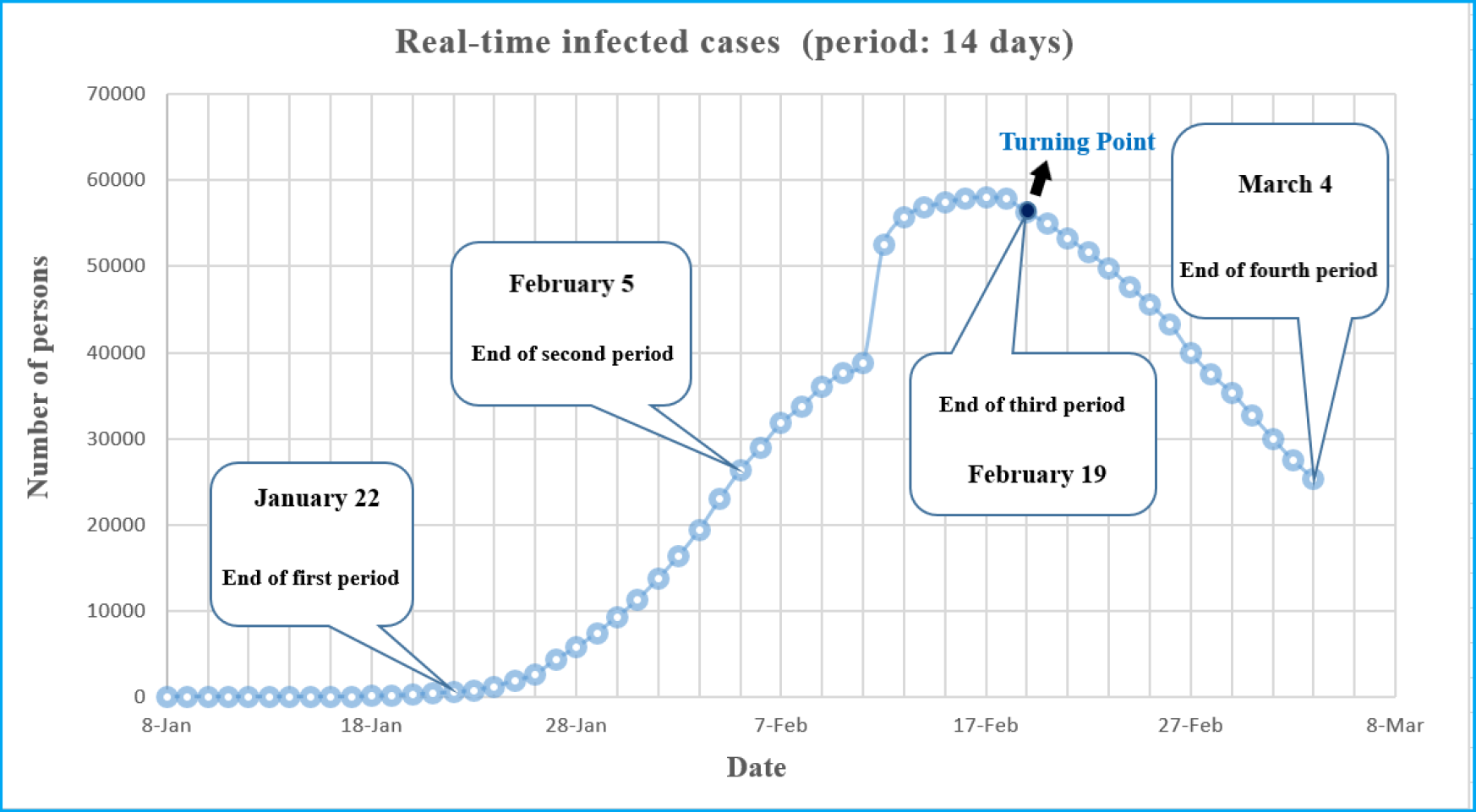
The number of real-time infected cases for each day = the number of accumulative infected cases for each day − the number of accumulative recovered cases for each day − the number of accumulative death cases for each day. The data comes from Figure 2.

To further test the validity of the reproduction numbers in Table 1, we substitute them into the SIR model (1)-(4) for simulating the evolution of China’s epidemic. To this end, let us first check the scope of application of the reproduction number formula (11), which is derived by equation (5). By the mean value theorem of integrals, equation (5) can be rewritten in the form:

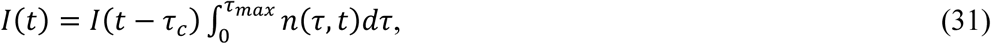

where 0 *≤ τ*_*c*_ *≤ τ*_*max*_.

By equation (6) and (31) we have:

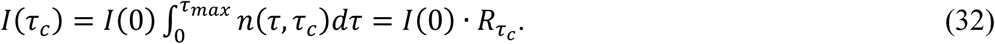

On the other hand, if we assume that *R*_*t*_ is a constant (step function) for each period, by equations (1)-(4) it is easy to get:

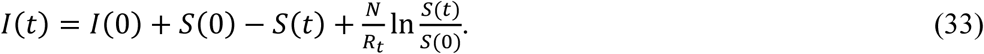

Let us order:

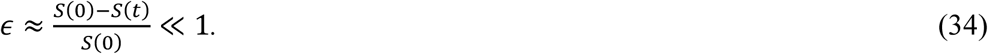

By using equation (34), equation (33) can be approximately written as:

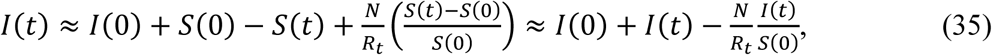

which implies

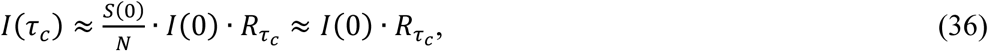

where equation (35) is derived by using the approximations |*R*_*t*_ − 1|. *R*(*t*) *≪ I*(*t*) and *S*(0) *≈ N*.

Comparing equations (32) and (36), we find that the reproduction number formula (11) can be applied to the SIR model (1)-(4) if the approximation (34) holds. Equation (34) implies (*N* − *S*(*t*))*/N ≪* 1. The approximation obviously holds for China, where *N ≈* 1.4 × 10^9^ and *N* − *S*(*t*) *≈* 1 × 10^5^. Therefore, we substitute the reproduction numbers in Table 1 into the SIR model (1)-(4) to simulate the evolution of China’s epidemic. The result is shown by Figure 5, where the evolutionary track (red circles) of the epidemic is well in accordance with that (black circles) of the real incident cases in China. The simulation result requires *τ ≈* 8, which agrees with our previous setting *τ* = 8.4 ± 3.8 [9]. Furthermore, we find that the reproduction numbers of quarantine periods in Table 1 can be fitted by an exponential function with *R*^2^ = 0.9924, see Figure 4. Therefore, we apply this exponential function to predicting the reproduction numbers for the next seven periods (from March 5, 2020 to June 10, 2020). The results have been listed in Table 2, where we also present the predicted values of the number of real-time infected cases for the last day in each period. These predicted values imply that China’s epidemic will gradually tend to disappear by May 2020, see the blue circles in Figure 5.

**Table 2:** Predicted values (2020)

| Period number $a$ | Period interval | $R_t$ | infected case number |
| --- | --- | --- | --- |
| 5 | 5-Mar to 18-Mar | 0.2545 | 8047 (last day) |
| 6 | 19-Mar to 1-Apr | 0.1119 | 1519 (last day) |
| 7 | 2-Apr to 15-Apr | 0.0492 | 253 (last day) |
| 8 | 16-Apr to 29-Apr | 0.0216 | 40 (last day) |
| 9 | 30-Apr to 13-May | 0.0095 | 6 (last day) |
| 10 | 14-May to 27-May | 0.0042 | 1 (last day) |
| 11 | 28-May to 10-Jun | 0.0018 | 0 (June 1) |

**Figure 4.**
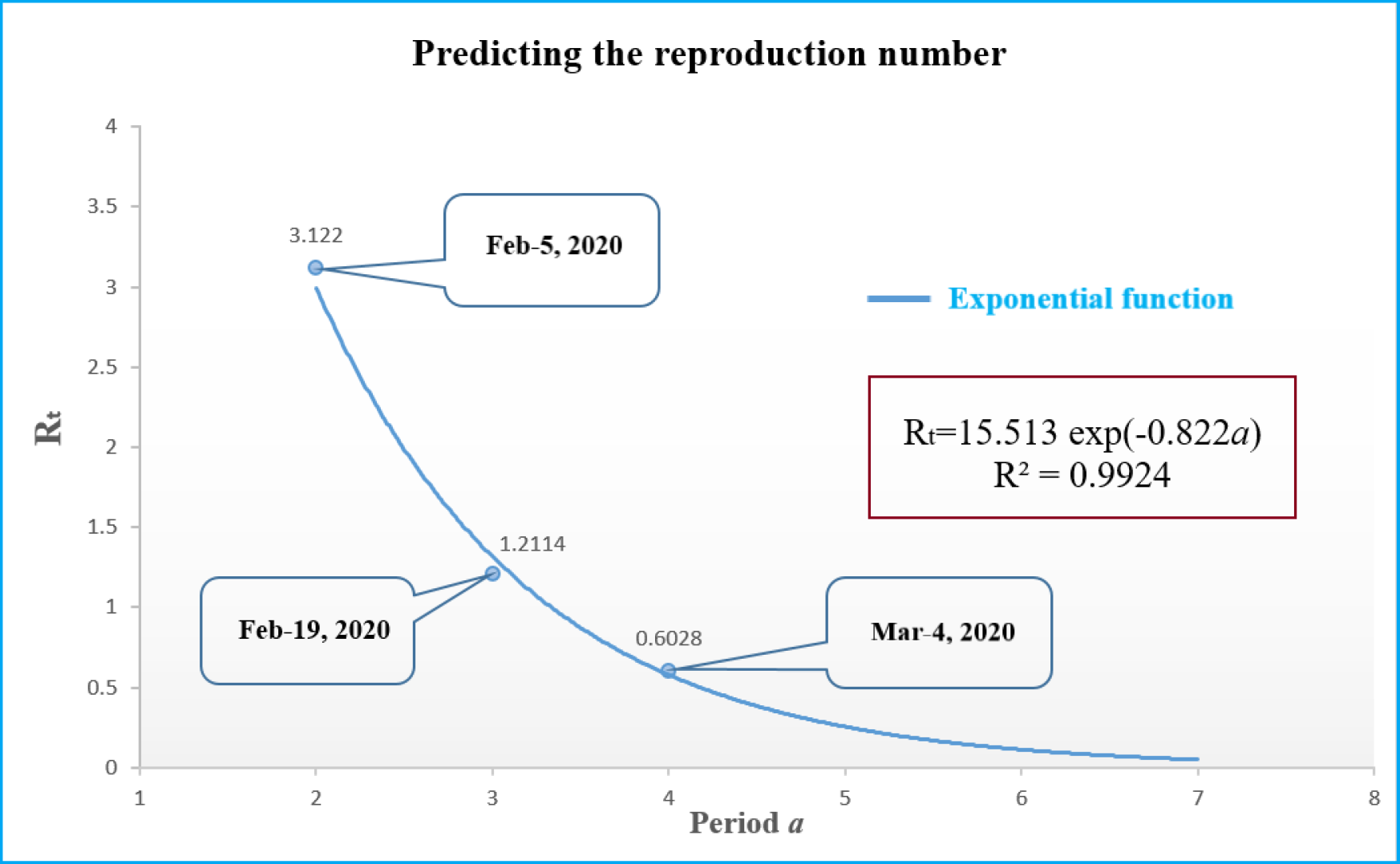
The reproduction numbers of quarantine periods in Table 1 are fitted by an exponential function.

**Figure 5.**
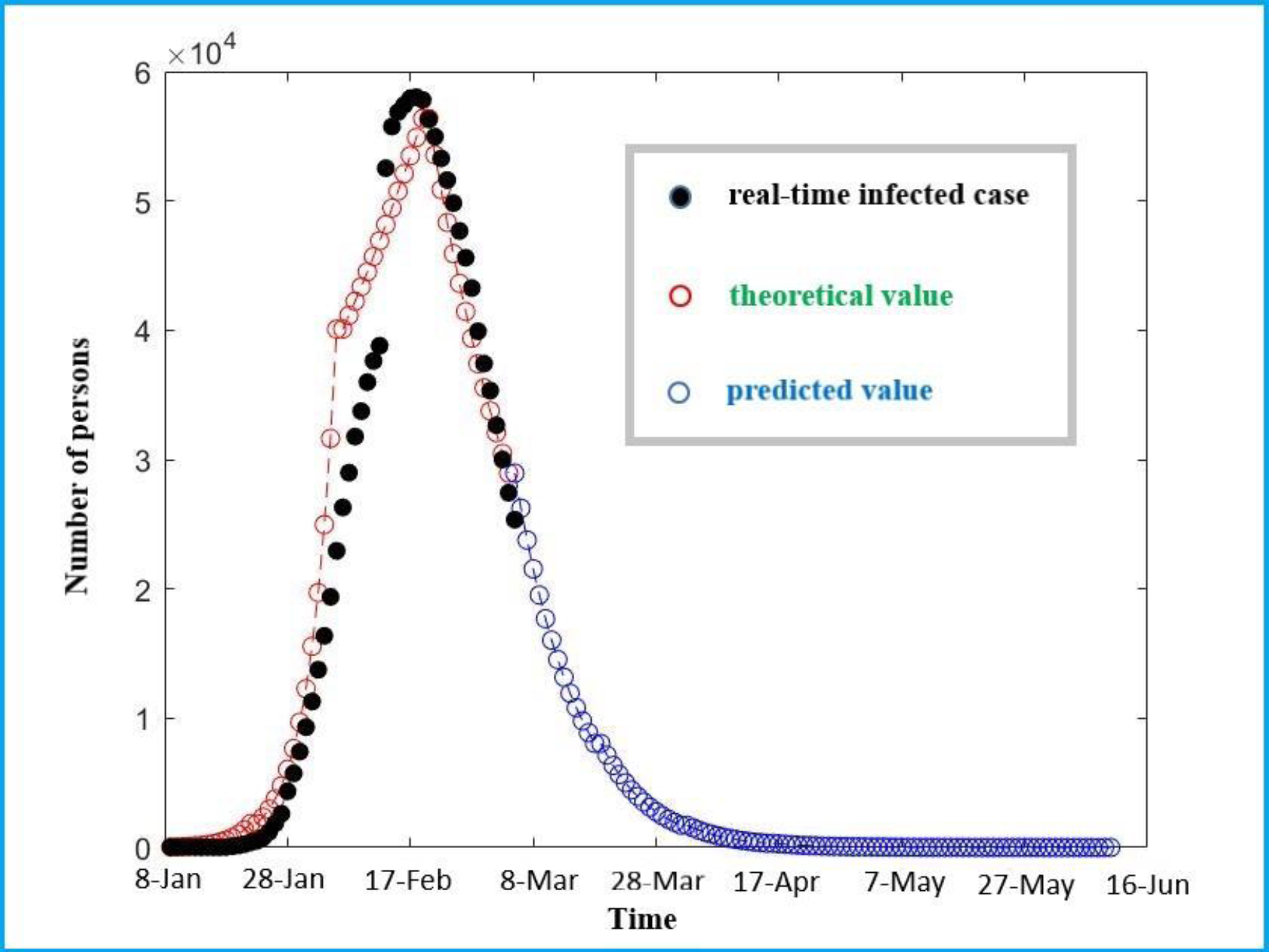
The SIR simulation result by using the reproduction numbers in Table 1 is showed by red circles and the SIR simulation result by using the reproduction numbers in Table 2 is showed by blue circles, where *N ≈* 1.4 × 10^9^ and *τ ≈* 8. The real-time infected cases in Figure 3 are showed by black circles.

In conclusion, to estimate the reproduction number, the probability distribution function of the generation interval of an infectious disease is required to be available; however, this distribution is often unknown. In the existing literature, many scholars used exponential distribution, normal distribution, Weibull distribution, and Gamma distribution to approximate the generation interval distribution. To do so, one needs to collect enough information about symptom onsets of all cases, which requires examining a large number of cases. By contrast, the maximum entropy method has more advantage of predicting probability distributions given the incomplete information. In this letter, we argue that, given the mean value and variance of the generation interval, one can determine its probability distribution function by using maximum entropy method. Because the overall data (population) of the generation interval is always absent, the maximum entropy method is a more convenient approach for estimating the probability distribution function of generation interval. By the maximum entropy method we first determine the probability distribution function of generation interval of COVID-19 and further apply it to estimating the real-time values of reproduction numbers of China’s epidemic. Plugging these estimated reproduction numbers into the susceptible-infectious-removed epidemic model, we simulate the evolutionary track of the epidemic in China, which is well in accordance with that of the real incident cases.

## Data Availability

I confirm the availability of all data in the paper.

